# Monoclonal antibody treatment drives rapid culture conversion in SARS-CoV-2 infection

**DOI:** 10.1101/2021.12.25.21268211

**Authors:** Julie Boucau, Kara W. Chew, Manish Choudhary, Rinki Deo, James Regan, James P. Flynn, Charles R. Crain, Michael D. Hughes, Justin Ritz, Carlee Moser, Joan A. Dragavon, Arzhang C. Javan, Ajay Nirula, Paul Klekotka, Alexander L. Greninger, Robert W. Coombs, William A Fischer, Eric S. Daar, David A. Wohl, Joseph J. Eron, Judith S. Currier, Davey M Smith, Jonathan Z. Li, Amy K. Barczak, for the ACTIV-2/A5401 Study Team

**Affiliations:** The Ragon Institute of MGH, MIT and Harvard, Cambridge, MA; University of California, Los Angeles, Los Angeles, CA; Brigham and Women’s Hospital, Harvard Medical School, Boston, MA; Harvard T.H. Chan School of Public Health, Boston, MA; University of Washington, Seattle, WA; National Institutes of Health, Bethesda, MD; Lilly Research Laboratories, San Diego, CA; University of North Carolina, Chapel Hill, N; Lundquist Institute at Harbor-UCLA Medical Center, Torrance, CA; University of California, San Diego, San Diego, CA; Massachusetts General Hospital, Harvard Medical School, Boston MA

## Abstract

Monoclonal antibodies (mAbs) are the treatment of choice for high-risk ambulatory persons with mild to moderate COVID-19. We studied viral culture dynamics post-treatment in a subset of participants receiving the mAb bamlanivimab in the ACTIV-2 trial. Viral load by qPCR and viral culture were performed from anterior nasal swabs collected on study days 0 (day of treatment), 1, 2, 3, and 7. Treatment with mAb resulted in rapid clearance of culturable virus in participants without treatment-emergent resistance. One day after treatment, 0 of 28 (0%) participants receiving mAb and 16 of 39 (41%) receiving placebo still had culturable virus (p <0.0001); nasal viral loads were only modestly lower in the mAb-treated group at days 2 and 3. Recrudescence of culturable virus was detected in three participants with emerging mAb resistance and viral load rebound. The rapid reduction in shedding of viable SARS-CoV-2 after mAb treatment highlights the potential role of mAbs in preventing disease transmission.

---

As the COVID-19 pandemic progresses, interventions have been developed to prevent transmission and progression to severe disease in infected persons. Monoclonal antibodies (mAbs) are currently first-line therapy for the outpatient management of high-risk individuals with mild to moderate COVID-19 (https://www.covid19treatmentguidelines.nih.gov). These mAbs have been shown to accelerate the decay of SARS-CoV-2 levels in the upper respiratory tract ^1,2^, but their effects on duration of shedding viable virus is unknown. While viral RNA is commonly used to assess viral burden, culture of viable virus from infected persons could be a more sensitive indicator of antiviral activity and potential for viral transmission. We hypothesized that reduction in shedding of viable virus might occur more rapidly than reduction in anterior nasal SARS CoV-2 RNA levels following mAb treatment. A full understanding of the potential benefits of mAbs and other treatments should help determine their optimal use for preventing and treating SARS-CoV-2 infection.

Bamlanivimab is a neutralizing mAb that binds to the spike protein of SARS-CoV-2, preventing uptake into host cells ^3^. It currently has emergency use authorization for use in conjunction with etesevimab for treatment of non-hospitalized, high-risk individuals with SARS-CoV-2 infection and for post-exposure prophylaxis. We performed viral culture analysis of participants enrolled in the ACTIV-2 randomized placebo-controlled trial of bamvalinimab monotherapy for non-hospitalized adults with mild to moderate COVID-19 ^4^. In that study, bamlanivimab treatment reduced respiratory tract (nasopharyngeal) viral load by 3 days post-treatment.

To compare shedding of viable virus and change in anterior nasal sample SARS CoV-2 RNA over time after treatment with mAb, we cultured virus from anterior nasal swabs collected from participants enrolled in the ACTIV-2 study^4^ who had baseline (pre-treatment, day 0) viral load of ⩾6 log_10_ SARS-CoV-2 RNA copies/mL and available swab samples from study days 0, 1, 2, 3 and 7. Participants with evidence of bamlanivimab resistance mutations at baseline or during follow-up based on our previous viral sequencing work ^5^ were initially excluded. Of the 317 participants in the ACTIV-2 study, 69 met inclusion criteria for the primary analysis in this study: 310 had available day 0 AN swabs, 94 had baseline viral load ⩾6 log_10_ SARS-CoV-2 RNA copies/mL, and 73 had swabs available at days 0, 1, 2, 3, 7. Four participants were excluded from the primary analysis due to emergent resistance identified in our previous work ^5^. Of the 69 participants meeting inclusion criteria, 39 participants fell into the placebo arm and 30 participants fell into the bamlanivimab arm (20 received the 7000mg dose and 10 received the 700mg dose). Baseline participant characteristics, including age, race, comorbidities, days of symptoms before enrollment, and serostatus, were similar between groups (Supp. Table 1). Baseline anterior nasal viral load was also similar between groups (Fig. 1A). Baseline viral culturability, as determined by cytopathic effect (CPE), was also similar between groups, with 39/39 (100%) participants in the placebo arm and 28/30 (93%) participants in the mAb arm with culture positive baseline sample (Fig. 1A). For samples with a sufficient number of positive wells to calculate semiquantitative viral culture titer (tissue culture infectious dose 50 [TCID_50_]) (34 placebo and 23 mAb samples), the relationship between SARS CoV-2 RNA and semiquantitative viral TCID_50_ was also similar between groups at the time of enrollment (Fig. 1B).

**Figure 1.**
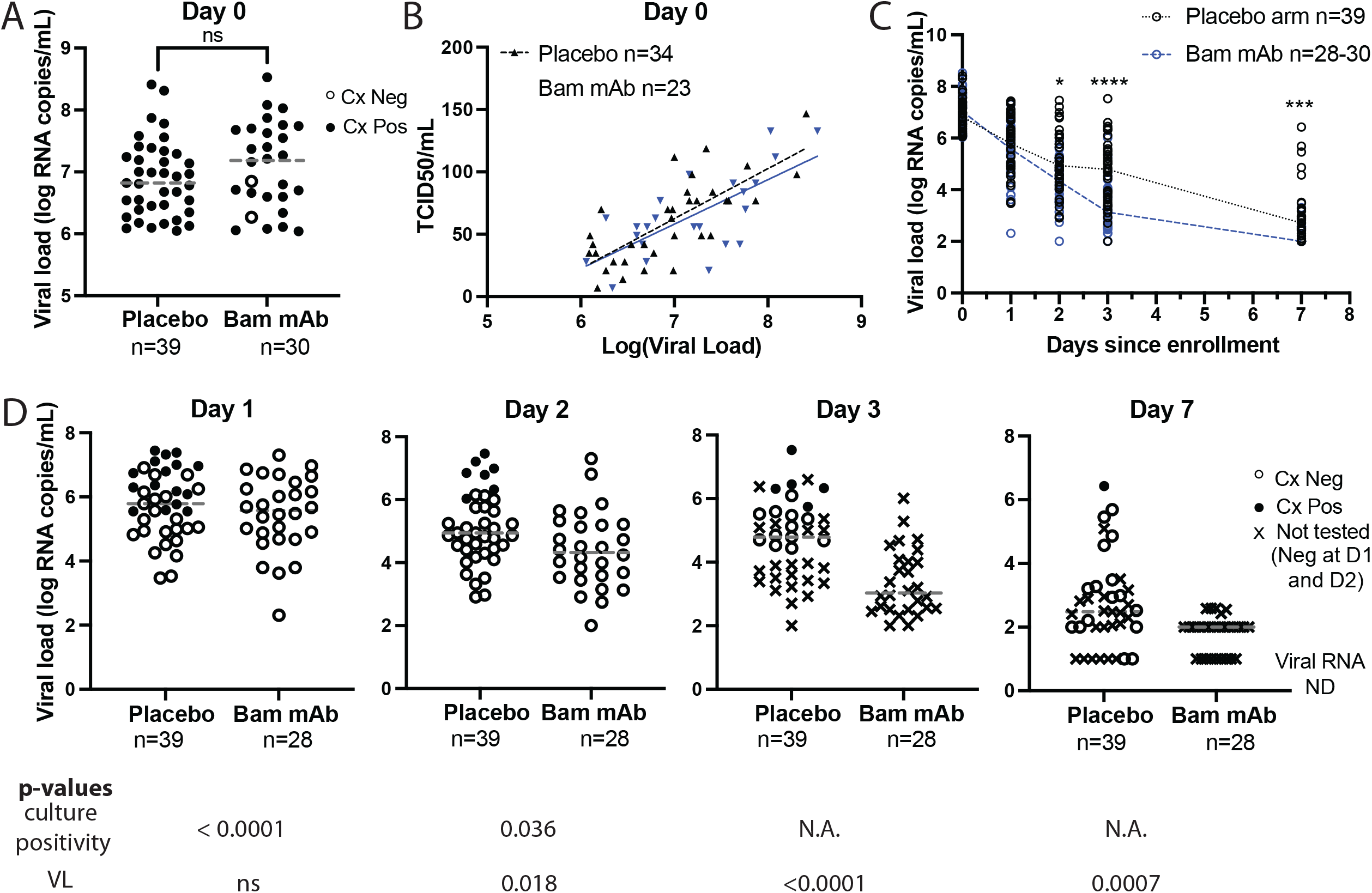
Bamlanivimab treatment results in rapid SARS-CoV-2 culture conversion. (A) Pre-treatment culture positivity and viral load (B) Pre-treatment TCID_50_ values vs. viral load; TCID_50_ could only be calculated for participants with ≥ 3 wells showing CPE. Spearman correlations: placebo r = 0.8482, p-value < 0.0001; Bam mAb r = .6365, p-value = 0.0011. (C) Decay in qPCR-determined viral load over time post-treatment (D) Culture positivity and viral load over time post-treatment. Cx, culture. Bam mAb, bamlanivimab monoclonal antibody.

Participants received either placebo or bamlanivimab on day 0. Anterior nasal sample SARS CoV-2 RNA was assessed prior to treatment (day 0) and at study days 1, 2, 3, and 7 post-treatment. Shedding of culturable virus was assessed prior to treatment (day 0) and at study days 1 and 2 post-treatment. For participants positive at either day 1 or day 2, culturability was further assessed at study days 3 and 7. While study day 1 nasal swab viral loads were similar between arms (Fig. 1C), significant difference in culture positivity was observed by day 1. In the placebo arm, culture positivity rate was 16/39 (41%) in the placebo arm vs 0/28 (0%) in the bamlanivimab arm (P<0.0001, Fig. 1D). In the placebo arm on day 1, the lowest viral load associated with a positive culture was 5.5 log_10_ RNA copies/mL, and 16/25 (64%) of placebo arm samples with a viral load ≥5.5 log_10_ RNA copies/mL were also found to be culture positive. In contrast, all 18 samples from the bamlanivimab arm with viral loads ≥5.5 log_10_ RNA copies/mL were culture negative. By day 2 post-treatment, 7 of 39 (18%) participants in the placebo arm were still culture positive; all participants in the bamlanivimab arm remained culture negative. 15 participants in the placebo arm and 0 participants in the bamlanivimab arm were culture positive at study day 1 and/or 2 and underwent additional testing on samples from days 3 and 7. Day 3, five of 15 tested placebo participants remained culture positive, and on study day 7, one of 15 tested placebo participants remained culture positive.

Viral resistance to bamlanivimab monotherapy has been described both *in vitro* and clinically, with resistance attributed to defined mutations in the SARS-CoV-2 spike protein ^5-7^. We previously identified participants from ACTIV-2 with emergent bamlanivimab resistance mutations ^5^ and these individuals were excluded from our primary analysis. We hypothesized that early viral culture clearance observed following bamlanivimab treatment was due to mAb binding and neutralization of virions, and that mAb resistance emergence would lead to renewed shedding of culturable virus. To test this hypothesis, we evaluated four participants with treatment-emergent bamlanivimab resistance mutations identified in our previous work ^5^; virus from three participants had emergent E484K mutation and one had emergent E484Q mutation. All four participants had positive viral cultures at day 0; similar to other participants in the bamlanivimab treatment arm of this substudy, all four individuals converted their cultures to negative on day 1 (Fig. 2A). However, the emergence of the E484K mutation following bamlanivimab treatment in the three participants was associated with rebound in viral loads and return of positive viral cultures with rising TCID_50_ levels (Fig. 2B-D). The participant with emergent E484Q mutation in their infecting virus had only modest increases in viral load and no re-emergence of positive viral cultures (Fig. 2E).

**Figure 2.**
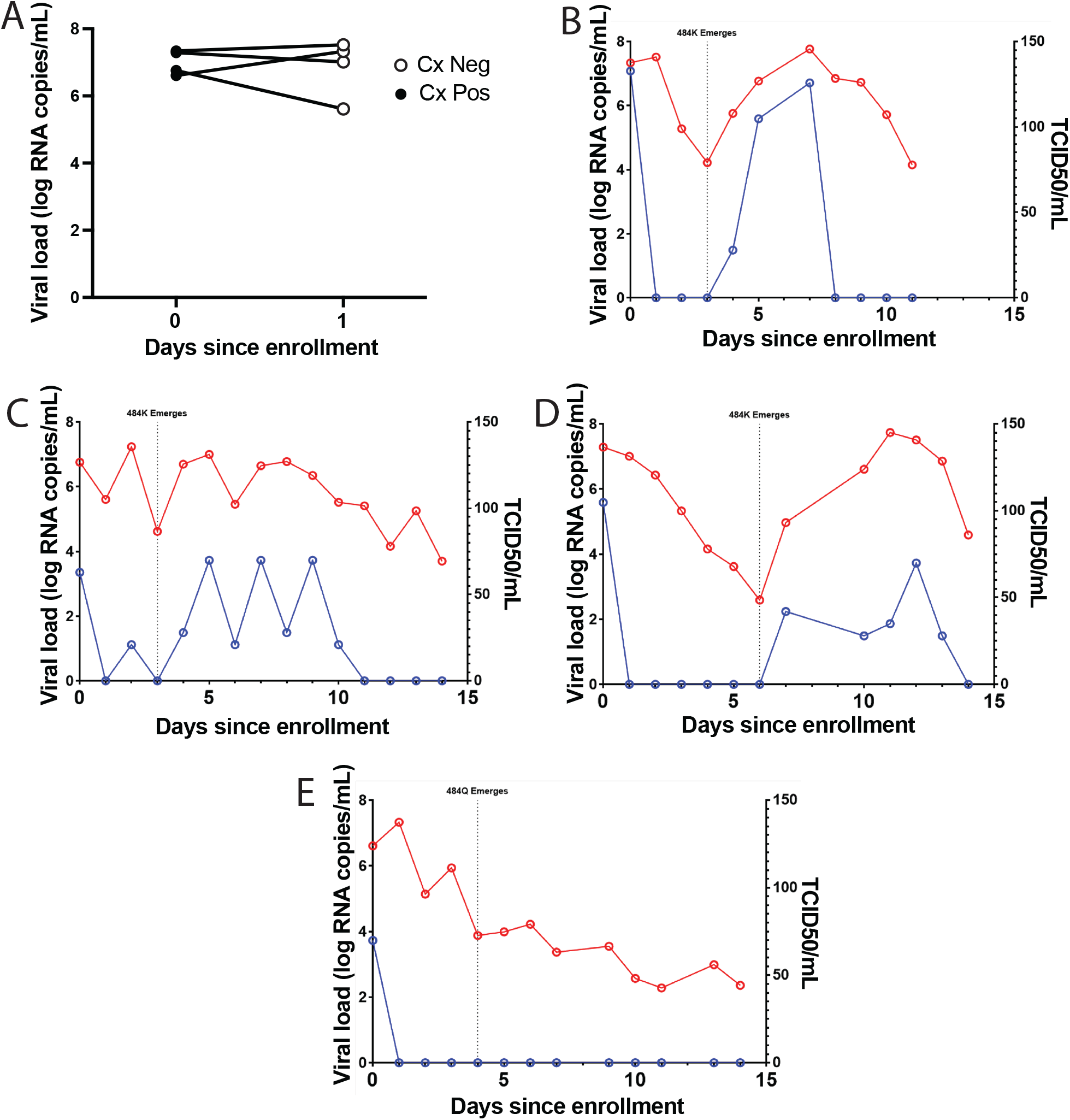
Emergence of bamlanivimab resistance mutations correlates with recrudescent shedding of culturable virus. (A) Viral load and culture positivity at baseline and day 1 post-treatment for four study participants with recrudescent shedding of culturable virus. Cx, culture (B-E) Viral load and TCID_50_ for four study participants whose infecting virus developed E484 mutations of the spike protein following bamlanivimab monotherapy.

Our results demonstrate that bamlanivimab treatment drives a rapid reduction in anterior nasal shedding of culturable virus that precedes a detectable reduction in viral load. Our findings suggest that viral culture assays could provide a means for the rapid evaluation of new mAb therapies in neutralizing infectious virus, which could accelerate the evaluation of mAbs in early phase clinical trials. While the effectiveness of a therapy is typically evaluated based on benefit to the infected individual, our results suggest that mAb treatment for COVID-19 may have an additional public health benefit, potentially reducing the period of infectiousness and consequently reducing the risk of secondary transmission. Whether this reduction in shedding of viable virus is unique to treatment with mAbs or would be similarly observed with other treatments, such as antiviral drugs with different mechanisms of action, is incompletely known. A recent study demonstrating reduction in culturable SARS-CoV-2 by study day 3 in participants treated with molnupiravir relative to placebo suggests that this benefit is not specific to mAb treatment, but may hold across a range of COVID-19 therapies ^8^. The time frame to negative culture warrants further evaluation across treatment modalities.

In this substudy, we focused on the subset of individuals most likely to have positive baseline cultures, namely those with baseline viral load of ⩾6 log_10_ SARS-CoV-2 RNA copies/mL. Based on the association we have previously observed between SARS-CoV-2 anterior nasal RNA level and likelihood of shedding culturable virus ^9-11^, individuals with lower baseline viral loads would be more likely to be culture negative at presentation. The precise relationship between baseline anterior nasal swab culture positivity, RNA level, and viral transmissibility remains to be determined; whether with further optimization, virus could be reliably cultured from samples with lower viral loads remains to be determined. However, the potential benefits of rapid culture conversion and reduced transmission might be greatest for individuals with high viral loads and those presenting early in the course of illness, who are most likely to be culture positive ^12^.

Additionally, our results underscore the importance of using mAbs in combinations to avoid the emergence of and selection for viral resistance. The re-emergence of culturable virus in a subset of ACTIV-2 participants with treatment-emergent drug resistance suggests that the development of resistance mutations may lead to a return of infectiousness and the risk of transmission of mAb-resistant strains. These results suggest that time to initial culture conversion and sustained culture conversion provide complementary information, and merit consideration of inclusion as outcome metrics in future studies of SARS-CoV-2 therapies.

Understanding the virologic consequences of therapeutic interventions for SARS-CoV-2 infection is critical for informing both the development of optimized treatment regimens and public health recommendations after treatment.

## Data Availability

The authors confirm that all data underlying the findings are fully available. Due to ethical restrictions, study data are available upon request from with the written agreement of the AIDS Clinical Trials Group and the manufacturer of the investigational product.

## Figure Legends

**Supplementary Table 1**. Baseline characteristics of participants in the placebo and bamlanivimab (700mg and 7000mg) arms.

## On-line Methods

### Study participants

ACTIV-2 is a multi-center randomized, blinded placebo-controlled phase 2/3 platform trial in non-hospitalized adults^4^. ACTIV-2 participants were enrolled at 39 sites in the U.S. between August 19 and November 17 2020. ACTIV-2 focused on the safety and efficacy of monoclonal antibody (mAb) bamlanivimab infusion in non-hospitalized participants with positive SARS-CoV-2 antigen or nucleic acid test within 7 days and less than 10 days of COVID-19 symptoms. There were two cohorts with different dosages of bamlanivimab (700 and 7000mg) in ACTIV-2, however due to low numbers, the two intervention groups were analyzed together. The study protocol was approved by the Mass General Brigham IRB as a secondary use protocol. Serial anterior nasal (AN) samples were self-collected by participants daily between enrollment (day 0) and day 14.

### Participant selection-substudy

Inclusion criteria for this substudy included baseline (pre-treatment, day 0) AN viral load of ⩾6 log_10_ SARS-CoV-2 RNA copies/mL, available AN swab samples from study days 0, 1, 2, 3 and 7. Participants otherwise meeting criteria for inclusion but with evidence of bamlanivimab resistance mutations at baseline or during follow-up based on our previous viral sequencing work ^5^ were excluded from the primary analysis and were analyzed as a separate emergent resistance subset.

### Viral culture

Viral culture experiments were performed as previously reported in the BSL3 laboratory of the Ragon Institute of MGH, MIT, and Harvard ^9-11^. Briefly, Vero-E6 cells were maintained in Dulbecco’s Modified Eagle Medium (DMEM) supplemented with HEPES, Penicillin/Streptomycin, Glutamine, and 10% Fetal Bovine serum (FBS), detached using Trypsin-EDTA and seeded at 75,000 cells per well in 24-well plates or 20,000 cells per well in 96-well plates 16-20 hours before infection. AN specimens were thawed on ice, filtered through a Spin-X 0.45 or 0.65um centrifugal filters at 10,000 x g for 5min and diluted them 1:10 in DMEM supplemented with HEPES, Penicillin/Streptomycin and Glutamine. 100uL of the diluted solution was used to inoculate triplicate wells in a 24-well plate for large scale culture experiments. After 1 hour of incubation at 37°C and 5% CO_2_, the viral inoculum was removed and 1mL of DMEM supplemented with HEPES, Penicillin/Streptomycin and Glutamine and 2% FBS (D2+) was added to each well. For the TCID_50_ experiments, 25ul of the undiluted filtrate was added to four wells of a 96-well plate and serially diluted (1:5) in D2+ media containing 5ug/mL of polybrene. The 96-well plates were then spinfected for 1 hour at 2000 x g at 37°C. The SARS-CoV-2 isolate USA-WA1/2020 strain and DMEM supplemented with HEPES, Penicillin/Streptomycin and Glutamine were used as positive and negative controls, respectively. We observed viral culture plates at 3- and 7-days post-infection with a light microscope and documented wells showing cytopathic effect (CPE). TCID_50_ titers were calculated using the Spearman-Karber method. In previous work, we had found that our assay was highly likely to grow virus for samples with RNA levels ≥ 6 log_10_ RNA copies/mL ^9-11^.

### Culture positivity

Specimens were defined as culture positive if at least 1 out of 3 wells showed CPE in the 24-well culture experiments or 1 out of 24 wells showed CPE in the TCID_50_ experiments. Because of the calculation method for TCID_50_ titers, TCID_50_ titers could only be calculated for samples with ≥ 3 wells showing CPE. Specimens with no observable CPE in either 24 well or TCID_50_ experiments were defined as culture negative. The USA WA-1/2020 strain was used as a positive control in all experiments.

### Statistics/Calculations

Statistical analyses were performed using PRISM software v9.2.0. Comparison of viral loads between the placebo and bamlanivimab treatment arms at the different time points was performed using the non-parametric two-tailed Mann-Whitney test. Comparison of proportion of culture conversion between the placebo and bamlanivimab treatment arms at the different time points was performed using two-sided Fisher’s exact test on a 2×2 contingency table.

### Funding

This work was supported by the National Institute of Allergy and Infectious Diseases of the National Institutes of Health under Award Number 3UM1AI068636-14S2, UM1 AI068634, UM1 AI068636 and UM1 AI106701. The viral culture work was performed in the Ragon Institute BSL3 core, which is supported in part by the NIH-funded Harvard University Center for AIDS Research (P30 AI060354). The content is solely the responsibility of the authors and does not necessarily represent the official views of the National Institutes of Health. The viral culture work was performed in the Ragon Institute BSL3 core, which is supported in part by the NIH-funded Harvard University Center for AIDS Research (P30 AI060354).

### Trial Registration

ClinicalTrials.gov Identifier: NCT04518410

## Acknowledgements

The authors would like to thank the participants, site staff, site investigators, and the entire ACTIV-2/A5401 study team. The authors would additionally like to thank the Ragon BSL3 core staff.

---

**Table 1.** Characteristics of participants in this substudy.

|  | Placebo arm<br>n=39 | Bamlanivimab arm<br>n=30 |
| --- | --- | --- |
| Age - median<br>(range) | 50 (20-71) | 50.5 (25-73) |
| Race – White<br>n (%) | 33 (85%) | 29 (97%) |
| Ethnicity –<br>Hispanic or<br>Latino n (%) | 10 (26%) | 3 (10%) |
| Gender –<br>Female n (%) | 24 (62%) | 15 (50%) |
| Comorbidities<br>– At least one<br>n (%) | 12 (31%) | 15 (50%) |
| Days of<br>symptoms<br>before<br>enrollment -<br>median<br>(range) | 4 (1-10) | 5 (3-8) |
| Seropositive<br>at baseline<br>(%)<sup>1</sup> | 0 (0%) | 1 (3%) |

<sup>1</sup>4 placebo and 2 intervention arm participants had unknown serostatus at baseline

